# Distinct baseline functional profiles of peanut-reactive T cells associate with sustained unresponsiveness after oral immunotherapy

**DOI:** 10.64898/2026.01.14.26344129

**Authors:** Duan Ni, Gabriela Pinget, Brigitte Santner-Nanan, Catherine L. Lai, Laurence Macia, Dianne E. Campbell, Peter Hsu, Ralph Nanan

## Abstract

**Background:** Peanut allergies continuously present urging public health challenges. Oral immunotherapy (OIT) is an important treatment option for peanut allergies, but its effectiveness varies, in terms of inducing desensitization (DS) or achieving long-term sustained unresponsiveness (SU). Identifying biomarkers to predict OIT outcomes is thus of great translational interests.

**Methods:** We thoroughly analyzed data from the POISED trial and our in-house OPIA trial, with a particular focus on the peanut-reactive T cells, in an attempt to identify potential biomarkers at baseline before OIT to distinguish DS and SU outcomes.

**Results:** In both the POISED trial and OPIA trial, we found that functional profiles of peanut-reactive T cells at baseline before OIT, such as their type II T helper (Th2) cell cytokine productions, including IL-4, were associated with the DS versus SU outcomes after OIT cessation.

**Conclusions:** Baseline peanut-reactive T cell functional profiles might provide new possibilities for biomarker discovery to predict peanut allergy OIT outcomes.

---

Food allergies, particularly peanut allergies, remain a critical public health challenge. Oral immunotherapy (OIT) represents an important treatment option for peanut allergy. OIT can readily induce desensitization (DS) in individuals with peanut allergy while they remain on treatment [1], which is referred to as no clinical reactivity to peanut with continued peanut allergen ingestion. However, particularly in older children, OIT still falls short of reliably inducing sustained unresponsiveness (SU), the ability to tolerate peanut after OIT cessation and an avoidance period. In this context, the majority of studies focus on the timepoint of OIT conclusion. Therefore, the ability to predict SU before OIT commencement is paramount, as this will facilitate improved patient stratification prior to treatment.

Previously, based on a Phase 2 randomized, controlled peanut OIT trial, *Peanut Oral Immunotherapy: Safety, Efficacy, Discovery (POISED, NCT02103270)*, Kaushik *et al*. reported that before OIT commencement, CD8^+^ T cell differentiation status and antigen presenting cell activation status could predict outcomes of SU vs DS (no clinical reactivity *versus* allergic responses during food challenge after OIT withdrawal and avoidance period) [2]. Based on the same cohort, using single cell RNA-seq (scRNA-seq), Han *et al*. found that a lower type 2 helper T (Th2) cell-related gene signature in peanut-reactive (pr) CD4^+^ effector T (Teff) cells (CD4^+^CD40L^+^) at baseline was linked to a higher likelihood of developing SU [3].

Inspired by their findings, we endeavoured to characterize the functional profiles of pr CD4^+^ T cells for further insights towards their association with SU. We first revisited the data from the POISED trial [2] (Figure 1A). Reanalyses after excluding outliers found that, at baseline, individuals with food allergy who developed SU 13 weeks after 2 years of OIT had less IL-4- and more TGFβ-expressing CD69^+^ pr CD4^+^ Teff cells (CD4^+^CD40L^+^CD69^+^) than DS individuals. No difference was found for productions of IFNγ, IL-9, IL-10, and IL-17 (Figure 1B).

**Figure 1.**
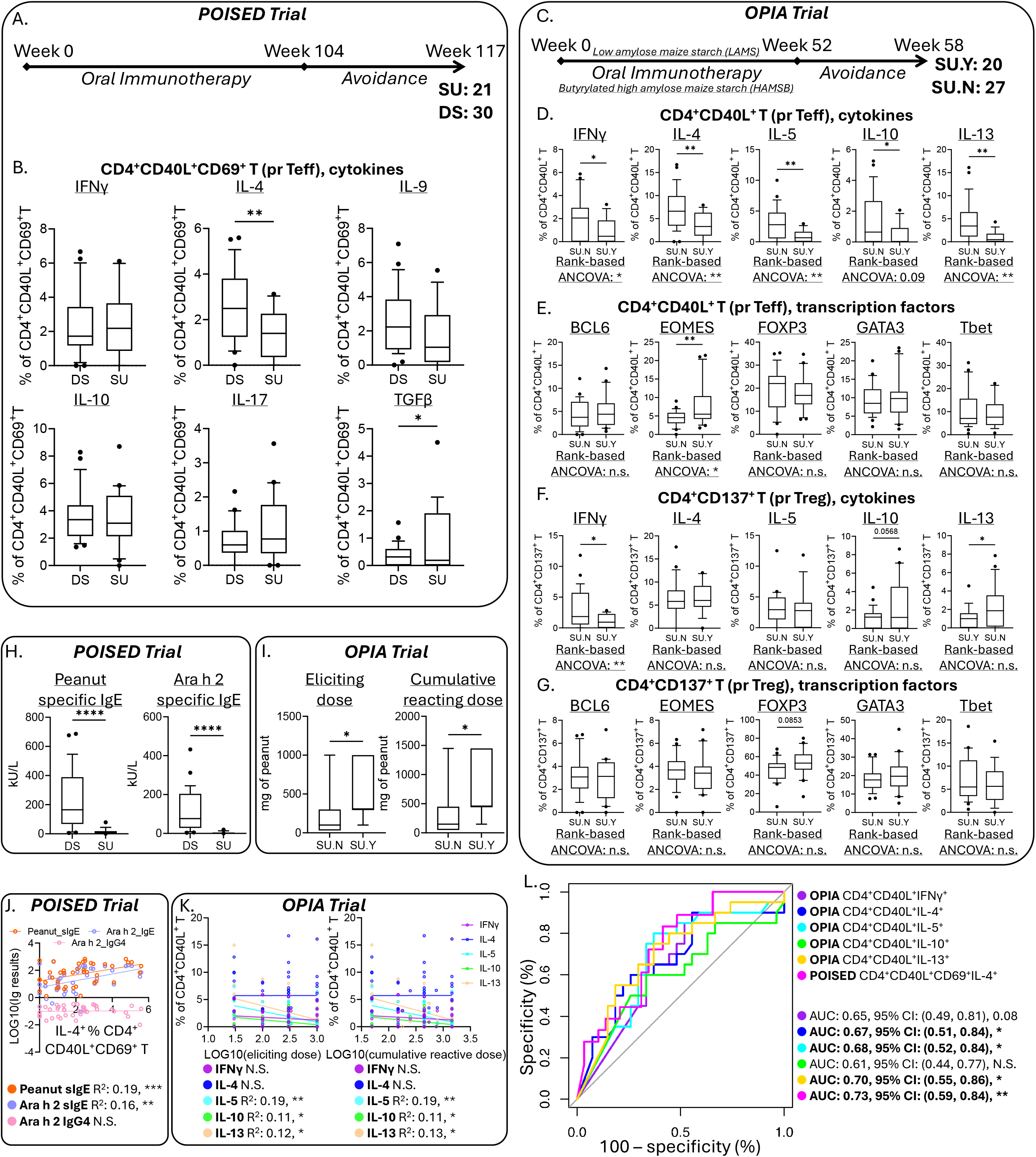
**A**. Overview of the timeline of Peanut Oral Immunotherapy: Safety, Efficacy, Discovery (POISED) clinical trial. **B**. Box charts showing the production of IFNγ, IL-4, IL-9, IL-10, IL-17 and TGFβ from CD69^+^ peanut reactive (pr) CD4^+^ (CD4^+^CD40L^+^CD69^+^) T effector (Teff) cells from individuals at baseline before oral immunotherapy (OIT), who later developed desensitization (DS) versus sustained unresponsiveness (SU) after OIT. **C**. Overview of the timeline of Oral Peanut Immunotherapy with Butyrate Adjuvant (OPIA) clinical trial. **D-E**. Box charts visualizing the cytokine (**D**) and transcription factor (**E**) profiles of peanut reactive (pr) CD4^+^ (CD4^+^CD40L^+^) T effector (Teff) cells from individuals at baseline before oral immunotherapy (OIT), who later developed (SU.Y) or failed (SU.N) sustained unresponsiveness after OIT. **F-G**. Box charts visualizing the cytokine (**F**) and transcription factor (**G**) profiles of peanut reactive (pr) CD4^+^ (CD4^+^CD137^+^) T regulatory (Treg) cells from individuals at baseline before oral immunotherapy (OIT), who later developed (SU.Y) or failed (SU.N) sustained unresponsiveness after OIT. Rank-based ANCOVA was run comparing SU.Y versus SU.N while adjusting for treatments with butyrylated high amylose maize starch (HAMSB) or low amylose maize starch (LAMS). **H**. Box charts depicting the peanut- and Ara h 2-specific IgE from individuals at baseline before oral immunotherapy (OIT), who later developed desensitization (DS) versus sustained unresponsiveness (SU) after OIT in the POISED trial. **I**. Box charts depicting the eliciting and cumulative reacting doses for peanuts for individuals at baseline before oral immunotherapy (OIT), who later developed (SU.Y) or failed (SU.N) sustained unresponsiveness after OIT in the OPIA trial. **J**. Correlation analyses in the POISED trial at baseline before OIT for IL-4 productions from CD69^+^ peanut reactive (pr) CD4^+^ (CD4^+^CD40L^+^CD69^+^) T effector (Teff) cells and log10-transformed peanut- and Ara h 2-specific IgE and Ara h 2-specific IgG_4_ levels. **K**. Correlation analyses in the OPIA trial at baseline before OIT for cytokine productions from pr CD4^+^ (CD4^+^CD40L^+^) Teff cells and log10-transformed eliciting and cumulative reacting peanut doses. **L**. Receiver operating characteristic (ROC) analysis for the prediction of SU outcomes in POISED and OPIA trials based on the baseline cytokine profiles of pr CD4^+^ Teff cells.

Next, similar comparative analyses were conducted using samples from our, double-blinded, randomized, placebo-controlled *Oral Peanut Immunotherapy with butyrate Adjuvant (OPIA, ACTRN12617000914369)* clinical trial [4] (Figure 1C). Here, we focused on the two arms that received peanut OIT and were supplemented with either butyrylated high amylose maize starch (HAMSB) adjuvant or placebo starch low amylose maize starch (LAMS) respectively. We previously reported that HAMSB and LAMS groups showed comparable performance in inducing SU 6 weeks after cessation of OIT [4]. We thus combined HAMSB and LAMS groups and comprehensively surveyed their pr T cell profile at baseline before OIT with high-dimensional spectral cytometry, assessing their potential associations with SU outcomes. As shown in Figure 1D, at baseline, pr CD4^+^ Teff cells (CD4^+^CD40L^+^) from the group of individuals who failed to develop SU (SU.N) produced more IFNγ, IL-4, IL-5, IL-10, and IL-13 than the cells from those who achieved SU (SU.Y). These differences generally remained significant even after adjusting for HAMSB/LAMS supplementation via rank-based analysis of covariance (ANCOVA). pr CD4^+^ Teff cells (CD4^+^CD40L^+^) from both groups expressed similar levels of transcription factors BCL6, FOXP3, GATA3 and Tbet, except EOMES, which was higher in the SU.Y group (Figure 1E). Furthermore, pr CD4^+^ regulatory T cells (Treg) (CD4^+^CD137^+^) were found to secrete more IFNγ in SU.N relative to SU.Y (Figure 1F), while showing no differences in other cytokines or transcription factors (Figure 1F-G).

Previously, it was reported that at baseline, higher peanut-specific and peanut allergen epitope-specific IgE levels were correlated with reduced success rates of SU post OIT [5]. Indeed, in the POISED cohort, we found that SU individuals had significantly lower levels of peanut-specific IgE and also IgE specific for Ara h 2 before OIT commencement (Figure 1H), while no difference was found for Ara h2-specific IgG_4_ (Figure S1). Similarly, in our OPIA trial, we also observed that the group who successfully developed SU (SU.Y) had higher baseline eliciting (ED) and cumulative reacting (CRD) doses of peanut compared to the SU.N group (Figure 1I). In these cohorts, baseline Th2 cytokine levels of pr CD4^+^ Teff cells were generally positively correlated with log10-transformed peanut- and Ara h 2-specific IgE concentrations (Figure 1J, POISED Trial) but negatively correlated with log10-transformed ED and CRD for peanut (Figure 1K, OPIA Trial).

Encouraged by these findings, receiver operating characteristic (ROC) analyses were carried out to predict the SU outcome based on the baseline Th2 cytokine profiles. As in Figure 1L, the IL-4 production from CD69^+^ pr CD4^+^ Teff cells (CD4^+^CD40L^+^CD69^+^) in POISED trial exhibited the best predictive performance (area under the curve (AUC) = 0.73, p = 0.008). In the OPIA study, levels of all Th2 cytokines secreted from pr CD4^+^ Teff cells (CD4^+^CD40L^+^) all had the potential to distinguish SU.Y versus SU.N groups (IL-4: AUC = 0.67, p = 0.04; IL-5: AUC = 0.68, p = 0.03; IL-13: AUC = 0.7, p = 0.02).

In summary, our cytometric study complements the scRNA-seq transcriptomic findings by Han *et al*., providing validation and extension at the protein level. These data support the notion that lower baseline Th2 profiles of peanut reactive (pr) CD4^+^ Teff (CD4^+^CD40L^+^) cells are linked to a higher likelihood of developing SU post peanut oral immunotherapy (OIT). Importantly, our findings were consistent across our OPIA trial and the POISED study, highlighting its general applicability. Moreover, we also provided additional insights by unveiling differences in the baseline pr CD4^+^ Treg (CD4^+^CD137^+^) cytokine profiles, and the transcription factor expression levels of both pr CD4^+^ Teff and pr CD4^+^ Treg cells in this regard.

In our OPIA trial dataset, in addition to lower levels of Th2 cytokines, we also observed lower Th1 cytokine (IFNγ) and IL-10 production in pr CD4^+^ Teff (CD4^+^CD40L^+^) cells from the group who developed SU, which were not found in either mass cytometry [2] or scRNA-seq [3] experiments based on the POISED cohort. This might be due to the differences in OIT time courses (OPIA: 1-year OIT, 6 weeks avoidance; POISED: 2-year OIT, 13 weeks avoidance) and peanut allergy patient demographics (e.g., OPIA: 10-16 years old, POISED: 7-55 years old), and the potential confounding effects of HAMSB/LAMS supplementations in the OPIA trial.

Nevertheless, despite these potential confounders, our results highlight the robust correlation between a Th2-biased pr CD4^+^ Teff (CD4^+^CD40L^+^) cell phenotype at baseline - before commencement of OIT - and SU. These findings might inform future patient stratification in food allergy and OIT treatment options. Our protein-based cytometric assays offer a more practical and cost-effective route for clinical translation than scRNA-seq experiments. Given the intermediate predictive power in our ROC analyses, future validations with larger cohorts are needed, and combining the cytokine profiles with other clinical parameters might further improve the predictive performance.

Importantly, in the OPIA study, despite differences in Th1 (IFNγ) and Th2 (IL-4, IL-5, and IL-13) cytokine profiles, similar levels of Th1 (Tbet) and Th2 (GATA3) signature transcription factors were found in pr CD4^+^ Teff (CD4^+^CD40L^+^) cells at baseline. Instead, more SU.Y pr CD4^+^ Teff (CD4^+^CD40L^+^) cells expressed EOMES. EOMES was previously reported to be a potential marker for human type 1 regulatory T cell (Tr1) [6]. Although our analyses found that both SU.Y pr CD4^+^ Teff (CD4^+^CD40L^+^) cells and pr CD4^+^ Treg (CD4^+^CD137^+^) produced less IFNγ, the detailed roles of EOMES in allergy, pr T cell, and particularly in SU development, warrant further investigation.

Similar to prior studies, our analyses have been limited by sample cellularity and availability to investigate other rare T cell subsets. These include follicular T helper cell (Tfh), and particularly pr CD8^+^ T cell, and CD8 Treg. Some investigations by others and us have shed light on the roles of CD8^+^ T cells in food allergy [4], which requires more exploration.

Collectively, our findings provide additional evidence linking a reduced baseline Th2-biased profile of pr CD4^+^ T effector cells to sustained unresponsiveness (SU) development after OIT, with potential relevance to novel biomarkers to predict OIT SU outcomes even before OIT commencement, pinpointing new possibilities for personalized food allergy treatments.

For all box charts, the boxes extend between 25^th^ to 75^th^ percentiles, while the whiskers mark 10^th^ to 90^th^ percentiles, and the lines in the middle denote the medians. (*: p<0.05, **: p<0.01, ***: p<0.0005, ****: p<0.0001, N.S.: not significant)

## Data Availability

All data produced in the present study are available upon reasonable request to the authors

## Disclosure statement

L.M. is now an employee of Sanofi-Aventis and this work was done while she was an employee of The University of Sydney. D.E.C. reports personal fees from DBV-Technologies and Westmead Fertility Centre, outside the submitted work. The others have nothing to declare.

The OPIA trial is funded by a National Health and Medical Research Council Australia Project Grant (NHMRC 1104134). D.N. was supported by the Norman Ernest Bequest Fund. C.L.L. is funded by the Centre for Food Allergy Research (CFAR) post-doctoral fellowship.

We acknowledge the contributions of the OPIA study group and help with data management by Katherine Thomson, Matthew Ward, and Ella Ward.

We thank the children and their families for participating in the OPIA trial.

## Ethics statement

The OPIA trial was registered on 22 June 2017 in Australian New Zealand Clinical Trials Registry as ACTRN12617000914369. The study was approved by the Human Research Ethics Committee of the Sydney Children’s Hospital Network (HREC/16/SCHN/372). Written informed consent was obtained from parents/guardians, and assent was obtained.

