## Supplementary Information for "Distinct baseline functional profiles of peanut-reactive T cells associate with sustained unresponsiveness after oral immunotherapy"

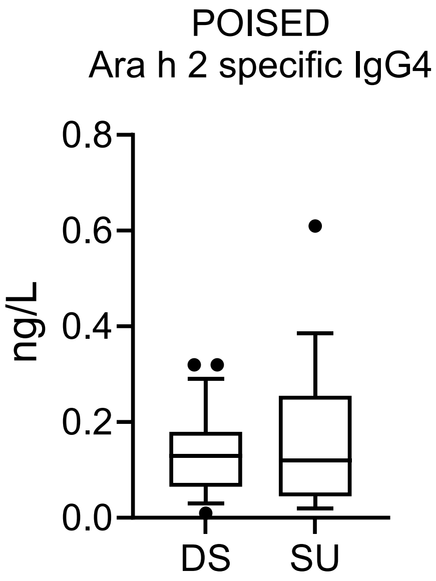


**Supplementary Figure S1.** Box charts depicting the Ara h 2-specific $IgG_4$ from individuals at baseline before oral immunotherapy (OIT), who later developed desensitization (DS) versus sustained unresponsiveness (SU) after OIT in the POISED trial.

### Materials and Methods

*Sex as a biological variable*

For both the POISED [1] and OPIA [2, 3] trials, both sexes were involved and they are generally balanced within the trial cohorts.

*Data curation for POISED trial*

Peanut Oral Immunotherapy: Safety, Efficacy, Discovery (POISED) study is a phase 2 randomized, controlled peanut OIT trial carried out by Kari Nadeau's team [1]. The overview of POISED study is depicted in Figure 1A and explained in its original publications [1]. Mass cytometry and serological (peanut- and Ara h 2-specific IgE, and Ara h 2-specific $IgG_4$) data were obtained from the prior related publication [4]. These data were first subject to interquartile range (IQR) analysis to exclude potential outliers and then analyzed with GraphPad PRISM.

*OPIA trial*

Oral Peanut Immunotherapy with Butyrate Adjuvant (OPIA) study was a single-centre randomized double-blinded placebo-controlled trial led by our team [2, 3]. It was registered on 22 June 2017 in the Australian New Zealand Clinical Trials Registry as ACTRN12617000914369. It was approved by the Human Research Ethics Committee of the Sydney Children's Hospital Network (HREC/16/SCHN/372). OPIA study's overall design is explained in Figure 2A and in our previous publications [2, 3]. In brief, the OPIA trial contained 3 arms: no OIT, 12-month OIT with butyrylated high amylose maize starch (HAMSB) as an adjuvant, and 12-month OIT with placebo low amylose maize starch (LAMS). For the purposes of this study, we focused on the 2 OIT treatment arms (HAMSB and LAMS) and combined them for comparisons both for t-test or for adjustment using rank-based analysis of covariance (ANCOVA).

*In-house spectral cytometry experiments based on OPIA trial*

Blood samples were collected from participants in the OPIA trial before OIT commencement. Peripheral blood mononuclear cells (PBMCs) were isolated, stimulated and stained as we previously reported [2, 3, 5]. In brief PBMCs were thawed and rested overnight in culture media (RPMI with 10% FBS), Penicillin/Streptomycin, and Glutamax (all from Thermo Fisher Scientific). For transcription factor analysis, cells were then cultured for 18 h in 96-well plates ($5 \times 10^6$ cells in 200 μL/well) in the presence of 2 μg/mL of anti-CD28, 1 μg/mL of anti-CD40, and 350 μg/mL of crude peanut extract (CPE). For cytokine analysis, cells were cultured for 6 h in 96-

well plates ($5 \times 10^6$ cells in 200 µL/well) in the presence of GolgiStop (BD, as per manufacturer's instructions), 2 µg/mL of anti-CD28, 1 µg/mL of anti-CD40, and 350 µg/mL of crude peanut extract (CPE). Endotoxin was removed using the Pierce endotoxin removal resin (Thermo Fisher Scientific). The final endotoxin concentration reached was < 5 EU/mL. Cells were then harvested for surface and intracellular staining as previously described [6] and acquired using the Cytek Aurora flow cytometer. Analysis was performed on FlowJo software (version 10.10.0).

*Statistics and ROC analysis*

Statistical analysis was run for each study cohort as described above.

Receiver operating characteristic (ROC) analysis was run in RStudio using the *pROC* (v1.18.0) package. We used different cytokine measurement results as predictors and attempted to model the OIT SU outcomes as responses.

**Study approval**

The OPIA trial was registered on 22 June 2017 in Australian New Zealand Clinical Trials Registry as ACTRN12617000914369. The study was approved by the Human Research Ethics Committee of the Sydney Children's Hospital Network (HREC/16/SCHN/372). Written informed consent was obtained from parents/guardians, and assent was obtained.

**Data availability**

Not applicable

**Acknowledgements**

The OPIA trial is funded by a National Health and Medical Research Council Australia Project Grant (NHMRC 1104134). D.N. was supported by the Norman Ernest Bequest Fund. C.L.L. is funded by the Centre for Food Allergy Research (CFAR) post-doctoral fellowship.

We acknowledge the contributions of the OPIA study group and help with data management by Katherine Thomson, Matthew Ward, and Ella Ward.

We thank the children and their families for participating in the OPIA trial.
